# Patient-centric radiology: Utilising large language models (LLMs) to improve patient communication and education

**DOI:** 10.64898/2026.02.23.26346923

**Authors:** Alexander Yip, Giles Craig, Javier Cortes-Ramirez, Nicole White, Kevin Shaw, Sandeep Reddy

## Abstract

**Purpose:** To evaluate whether large language models (LLMs) can enhance clinician-patient communication by simplifying radiology reports to improve patient readability and comprehension.

**Methods:** A randomised controlled trial was conducted at a single healthcare service for patients undergoing X-ray, ultrasound or computed tomography between May 2025 and June 2025. Participants were randomised in a 1:1 ratio to receive either (1) the formal radiology report only or (2) the formal radiology report and an LLM-simplified version. Readability scores, including the Simple Measure of Gobbledygook, Automated Readability Index, Flesch Reading Ease, and Flesch-Kincaid grade level, were calculated for both reports. Statistical analysis of patient readability and comprehension levels, factual accuracy and hallucination rates for LLMs was assessed using a combination of binary and 5-point Likert scales, open-ended survey questions, and independent review by two radiologists.

**Results:** 59/120 patients were randomised to receive both the formal and LLM-simplified radiology reports. Readability of LLM-simplified reports significantly improved with the reading level required for formal reports equivalent to a university-standard (11^th^-13^th^ grade) compared to a middle-school standard (5^th^-9^th^ grade) for simplified reports (rank biserial correlation=0.83, p<0.001). Patients with both reports demonstrated a significantly greater comprehension level, with 95% reporting an understanding level greater than 50%, compared with 46% without the simplified report (rank biserial correlation = 0.67, p < 0.001). All LLM-simplified reports were considered at least somewhat accurate with a minimal hallucination rate of 1.7%. Importantly, no hallucinations resulted in potential patient harm. 118/120 (98.3%) patients expressed interest in simplified radiology reports to be included in future clinical practice.

**Conclusion:** This study provides evidence that LLMs can simplify radiology reports to an accessible level of readability with minimal hallucination. LLMs improve both ease of readability and comprehension of radiology reports for patients. Therefore, the rapid advancement of LLMs shows strong potential in enhancing patient-radiologist communication as patient access to electronic health records is increasingly adopted.

**Highlights:**

- Radiology reports can be complex and difficult for patients to read and interpret
- Strong patient demand exists for simplified radiology reports
- Large language models (LLMs) such as GPT-4o show promise in simplifying radiology reports
- LLMs credibly simplify radiology reports with minimal hallucination rates
- LLMs improve both patient readability and comprehension of radiology reports

## Introduction

Patient access to electronic health records and online patient portals is increasingly adopted within healthcare settings, reflecting the global shift towards a patient-centred approach [1]. In Australia, recent legislative reform as part of the ‘Modernising My Health Record – Sharing by Default Bill 2024’ now mandates healthcare providers to share key health information, including radiology reports, with My Health Record (MHR) [2]. Whilst these legislative changes promote greater transparency and empower patients to take control of their health outcomes, there remains a clear discrepancy between the level of health literacy required to read and understand radiology reports.

Traditionally, radiology reports incorporate complex medical jargon intended for interpretation by referring clinicians. However, this is often inadvertently at the detriment of the patient due to their limited comprehension of medical terminology and may result in increased patient anxiety, confusion and misinterpretation of report findings [3, 4]. The average American demonstrates a reading level equivalent to the eighth grade, whilst 44% of Australians have a literacy level at or below the tenth grade [5, 6]. Both American and Australian guidelines therefore recommend written content to be ideally pitched at a readability level equivalent to or below the sixth and seventh grades, respectively. However, several studies demonstrate that, on average, radiology reports are written at a readability level equivalent to the twelfth to thirteenth grade, with only 4% of radiology reports readable at the eighth-grade level [7, 8]. Yet, effective communication between radiologists and patients remains crucial in improving patient experience, safety and health outcomes [9]. With the recent legislative changes, it is therefore paramount to communicate radiological report findings in a manner comprehensible to both healthcare providers and patients.

Numerous strategies have been proposed to improve the readability of radiology reports, primarily for clinicians, including standardised structured reporting, additional summary statements, annotated reports with hyperlinks to the radiology lexicon, anatomical illustrations and additional radiologist explanations of the report findings. Yet, few strategies have been designed for patients, and to our knowledge, none have been implemented in Australia. Also, the aforementioned strategies are often impractical in high-volume clinical settings due to significant time and workload constraints, as well as increased disruptions, which can lead to radiologist burnout in an ever-increasing workflow environment [4, 10-12].

Recent advances in artificial intelligence (AI), most notably large language models (LLMs) such as ChatGPT (OpenAI), demonstrate promising potential to transform the radiological landscape. LLMs are a subset of generative AI that utilise natural language processing to understand, interpret, and generate probabilistic, predictive language based on large numbers of pretrained parameters and data. The premise of LLMs relies on the revolutionary transformer architecture, which enables encoded tokens embedded within a vector database to be independently analysed in parallel whilst assigning an important weighting factor known as the attention mechanism [13]. Whilst various radiological applications of LLMs have been proposed, including automatic report generation, standardised structuring reporting, speech-to-text error recognition and report translation, there is a growing focus on radiology report summarisation as a means to bridge the apparent patient health literacy gap, limit the number of unnecessary patient-initiated enquires directed at clinicians and therefore, minimise radiologist workflow burden [14-22].

Numerous studies have demonstrated that LLMs can accurately synthesise radiology report findings, improving readability across a range of imaging modalities [18, 19, 21, 22]. To our knowledge, there is limited data on how patients read and comprehend LLM-simplified radiology reports in the context of their own health. Given the imminent legislative requirement that radiology reports be shared by default with MHR, there remains a pressing need to explore how AI-driven tools can be leveraged to improve health communication without compromising patient safety.

This study aims to assess whether LLMs can improve clinician-patient communication by accurately simplifying radiology reports to a patient-appropriate readability level, as compared with the formal radiology report. The primary outcome is the difference in the patient’s readability and understandability of the simplified radiology report compared with the formal radiology report. The secondary outcomes include assessing the factual accuracy of LLMs in simplifying radiology reports, quantifying hallucination rates and evaluating their potential to cause patient harm through increased anxiety or misunderstanding.

## Methods

### Patient population and participant recruitment

This study was conducted as a single-centre randomised controlled trial. Eligible participants were aged 18 years or older and undertook an outpatient medical imaging examination (X-ray, ultrasound or computed tomography (CT)) at Barwon Health (Victoria, Australia) between May 2025 and June 2025 **(Figure 1)**. An outpatient was defined as a patient who, at the time of attendance for a diagnostic imaging examination, was not admitted or receiving treatment within the hospital. Participants were excluded from the study if they were admitted to the hospital following imaging for urgent evaluation and/or treatment, had prior formal medical training, were unable to read English, had a new diagnosis of a sinister pathology or underwent an antenatal obstetric ultrasound. Patients requiring urgent evaluation were excluded as they would be expected to receive prompt explanations from treating clinicians, reducing the relevance of a simplified report. Individuals with prior medical training were omitted as medical practitioners are the intended target audience for radiology reports, and simplified reports would offer limited additional value. Non-English readers were excluded to avoid the confounding effects of interpreter and translation involvement. Newly diagnosed sinister pathologies, such as ectopic pregnancies or new cancer diagnoses, as determined by a radiologist, were excluded to minimise the risk of undue patient anxiety and distress. Obstetric ultrasound reports were excluded because they rely on graphical and numerical data that may not be meaningfully simplified.

**Figure 1:**
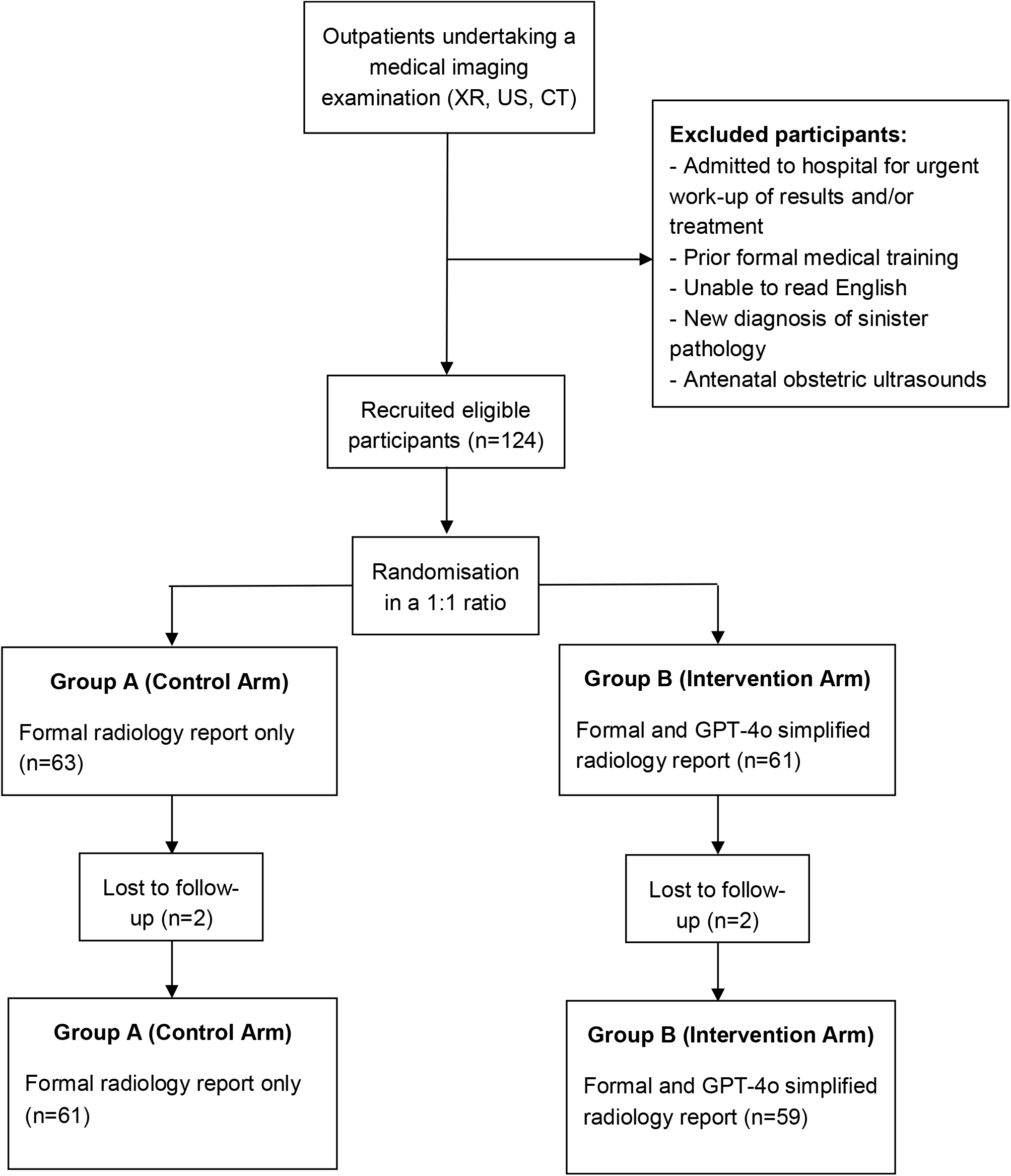
Flow diagram of the patient selection and randomisation process CT = Computed tomography; GPT = Generative Pre-trained Transformer; US = Ultrasound; XR = X-ray.

Participants were randomised in a 1:1 ratio to receive either (1) the formal radiology report only or (2) the formal radiology report and an LLM-simplified version.

### Ethics clearance

Institutional review board approval was obtained from the Barwon Health Human Research Ethics Committee (local reference number 25/48; approved on 2 May 2025) and the Clinical Informatics Governance Committee. Written consent was obtained from all recruited participants.

### Large language model

De-identified radiology reports were simplified to a readability level equivalent to the sixth-grade level using a single zero-shot prompt via Microsoft 365 CoPilot, an LLM based on the Generative Pretrained Transformer-4o (GPT-4o) architecture **(Supplementary material 1)**. Zero-shot prompting refers to the generation of an output response based on the LLM’s pre-trained knowledge rather than providing the LLM with prior examples to learn from. The LLM was hosted locally within the institutional information technology (IT) infrastructure to preserve the privacy and security of patient data.

### Data collection

The self-perceived readability and understandability of participants’ individual radiology reports were evaluated. Readability refers to how easy a radiology report is to read, based on linguistic characteristics such as sentence length, structure, and word complexity. Understandability was defined as participants’ ability to accurately interpret and comprehend the contents of the radiology report, including medical jargon and report findings.

Both objective and subjective readability metrics were calculated for the formal and GPT-4o simplified reports. In the absence of a universally accepted reference standard for evaluating readability, a combination of objective readability metrics, including the Simple Measure of Gobbledygook (SMOG) index, Automated Readability Index (ARI), Flesch Reading Ease and Flesch-Kincaid grade level scores, was employed to improve reliability. Each readability metric is calculated using a standardised formula derived from linguistic characteristics including word and sentence length, sentence structure and syllable count. Although these readability metrics yield quantitative scores, the values correlate with discrete educational grade levels, facilitating meaningful interpretation. A standardised questionnaire **(Supplementary material 2)** was administered to patients to evaluate the perceived readability and understandability of the formal and GPT-4o-simplified radiology reports, using a combination of five-point Likert scales and free-text responses. The patient’s readability of the report was measured on a scale of 5 (very easy, easy, neutral, difficult, and very difficult). The patient’s understanding of the report was measured with five levels (minimal (<25%), somewhat minimal (25-50%), approximately half (50%), somewhat more than half (50-75%), and almost all (>75%).

Two consultant radiologists (G.C. and K.S.) independently assessed the factual accuracy of GPT-4o by comparing the formal and LLM-simplified radiology reports using a five-point Likert scale. Factual accuracy was measured on a five-point scale (very inaccurate, inaccurate, somewhat accurate, accurate, and very accurate). The presence of hallucinations and their potential to cause patient harm in the form of increased anxiety or miscomprehension were also evaluated using a binary scale. Both radiologists were blinded from each other’s responses. Patient demographic data was collected using binary and free-text responses.

### Statistical analysis

The target sample size has been proposed based on the minimum improvement expected for the primary outcome, and feasibility with respect to participant recruitment. Previous research has estimated that approximately 20% of patients comprehend formal radiology reports [21]. If the intervention is effective, we expect at least 50% of patients to understand the LLM-simplified radiology report. Based on these estimates, a minimum of 104 participants (52 per arm) provides an expected power of 0.9. The expected power is based on a two-sample proportion test with a two-sided alternative hypothesis and a 5% significance level.

Descriptive statistics, expressed as proportions (categorical variables) and means (continuous variables), were used to characterise the sample. For the primary outcomes, a Mann-Whitney U test was used to compare the readability and understandability between the two groups (formal and LLM-simplified radiology reports), and a paired t-test was used to compare the readability indexes of the two groups. For the secondary outcomes, the X^2^ test was used to compare differences in the radiologists’ evaluation of factual accuracy and hallucination in the simplified report.

All analyses were performed in R (ver. 4.1) using the R Stats Package [23]. Values with p□<□0.05 were considered statistically significant.

## Results

A total of 120 patients who underwent either an X-ray, ultrasound or CT were included, with 59 participants receiving both the formal and GPT-4o simplified radiology reports. The average age was 54 years, with a larger proportion of females (67%). The characteristics of the studied population and the differences between groups are shown in **Table 1**.

**Table 1:**
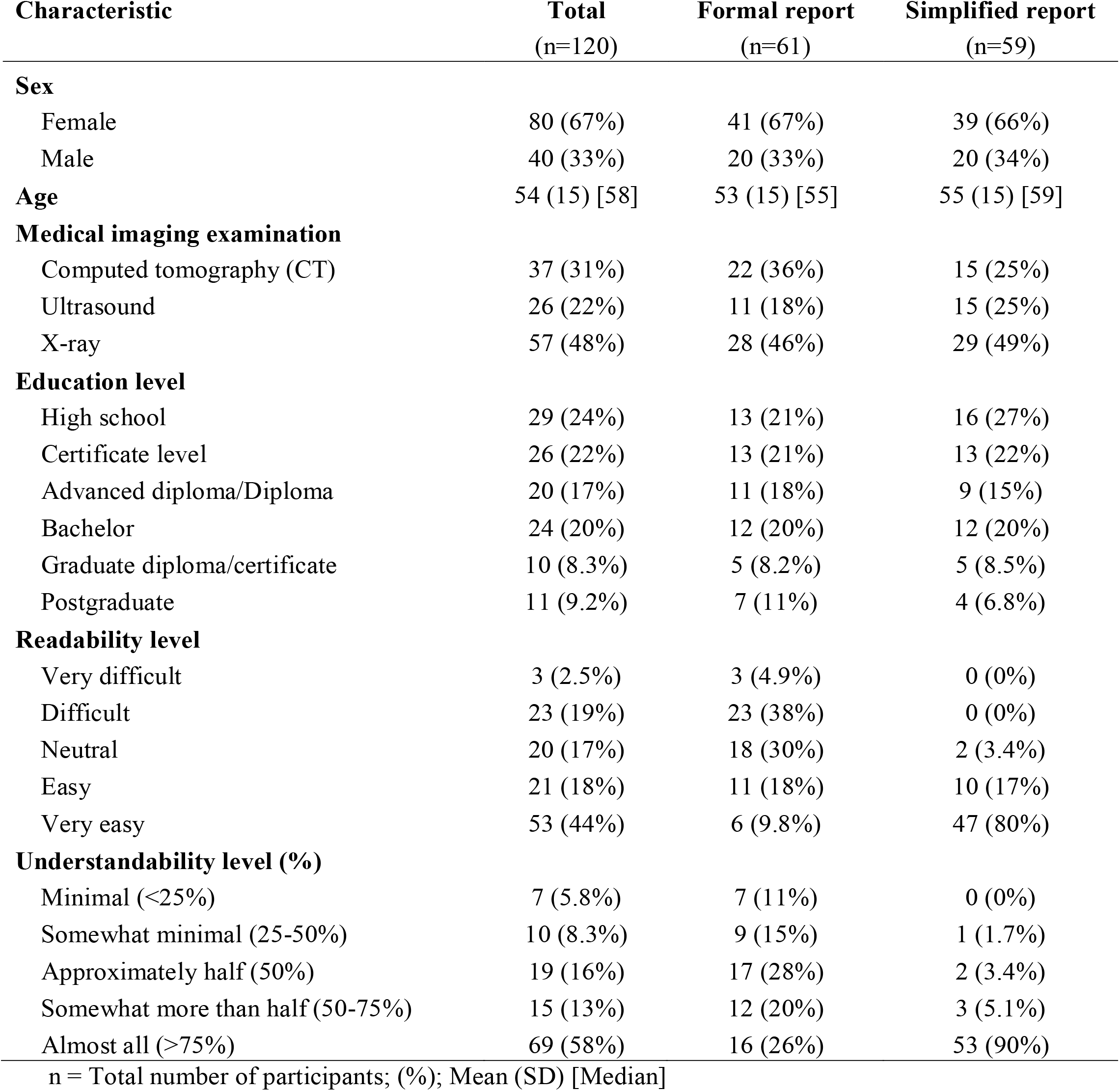
Characteristics of study participants by treatment group.

| **Characteristic** | **Total**<br>**(n=120)** | **Formal report**<br>**(n=61)** | **Simplified report**<br>**(n=59)** |
| --- | --- | --- | --- |
| **Sex** |  |  |  |
| Female | 80 (67%) | 41 (67%) | 39 (66%) |
| Male | 40 (33%) | 20 (33%) | 20 (34%) |
| **Age** | 54 (15) [58] | 53 (15) [55] | 55 (15) [59] |
| **Medical imaging examination** |  |  |  |
| Computed tomography (CT) | 37 (31%) | 22 (36%) | 15 (25%) |
| Ultrasound | 26 (22%) | 11 (18%) | 15 (25%) |
| X-ray | 57 (48%) | 28 (46%) | 29 (49%) |
| **Education level** |  |  |  |
| High school | 29 (24%) | 13 (21%) | 16 (27%) |
| Certificate level | 26 (22%) | 13 (21%) | 13 (22%) |
| Advanced diploma/Diploma | 20 (17%) | 11 (18%) | 9 (15%) |
| Bachelor | 24 (20%) | 12 (20%) | 12 (20%) |
| Graduate diploma/certificate | 10 (8.3%) | 5 (8.2%) | 5 (8.5%) |
| Postgraduate | 11 (9.2%) | 7 (11%) | 4 (6.8%) |
| **Readability level** |  |  |  |
| Very difficult | 3 (2.5%) | 3 (4.9%) | 0 (0%) |
| Difficult | 23 (19%) | 23 (38%) | 0 (0%) |
| Neutral | 20 (17%) | 18 (30%) | 2 (3.4%) |
| Easy | 21 (18%) | 11 (18%) | 10 (17%) |
| Very easy | 53 (44%) | 6 (9.8%) | 47 (80%) |
| **Understandability level (%)** |  |  |  |
| Minimal (<25%) | 7 (5.8%) | 7 (11%) | 0 (0%) |
| Somewhat minimal (25-50%) | 10 (8.3%) | 9 (15%) | 1 (1.7%) |
| Approximately half (50%) | 19 (16%) | 17 (28%) | 2 (3.4%) |
| Somewhat more than half (50-75%) | 15 (13%) | 12 (20%) | 3 (5.1%) |
| Almost all (>75%) | 69 (58%) | 16 (26%) | 53 (90%) |

n = Total number of participants; (%); Mean (SD) [Median]

### Primary outcomes

#### Readability scores

The GPT-4o simplified reports were subjectively assessed as either ‘easy’ or ‘very easy’ to read by 97% of patients, with no patients reporting them as ‘very difficult’ or ‘difficult’ to read **(Table 1, Figure 2a)**. Only 28% of patients who read the formal radiology report scored it as ‘easy’ or ‘very easy’, with 43% of patients scoring the formal report as either ‘very difficult’ or ‘difficult’ to read. There was a statistically significant difference in the rank-sum test for the GPT-4o simplified report compared with the formal report. This was associated with higher ranks for scores in the intervention arm, as indicated by a strong positive rank-biserial correlation **(Table 2)**.

**Figure 2:**
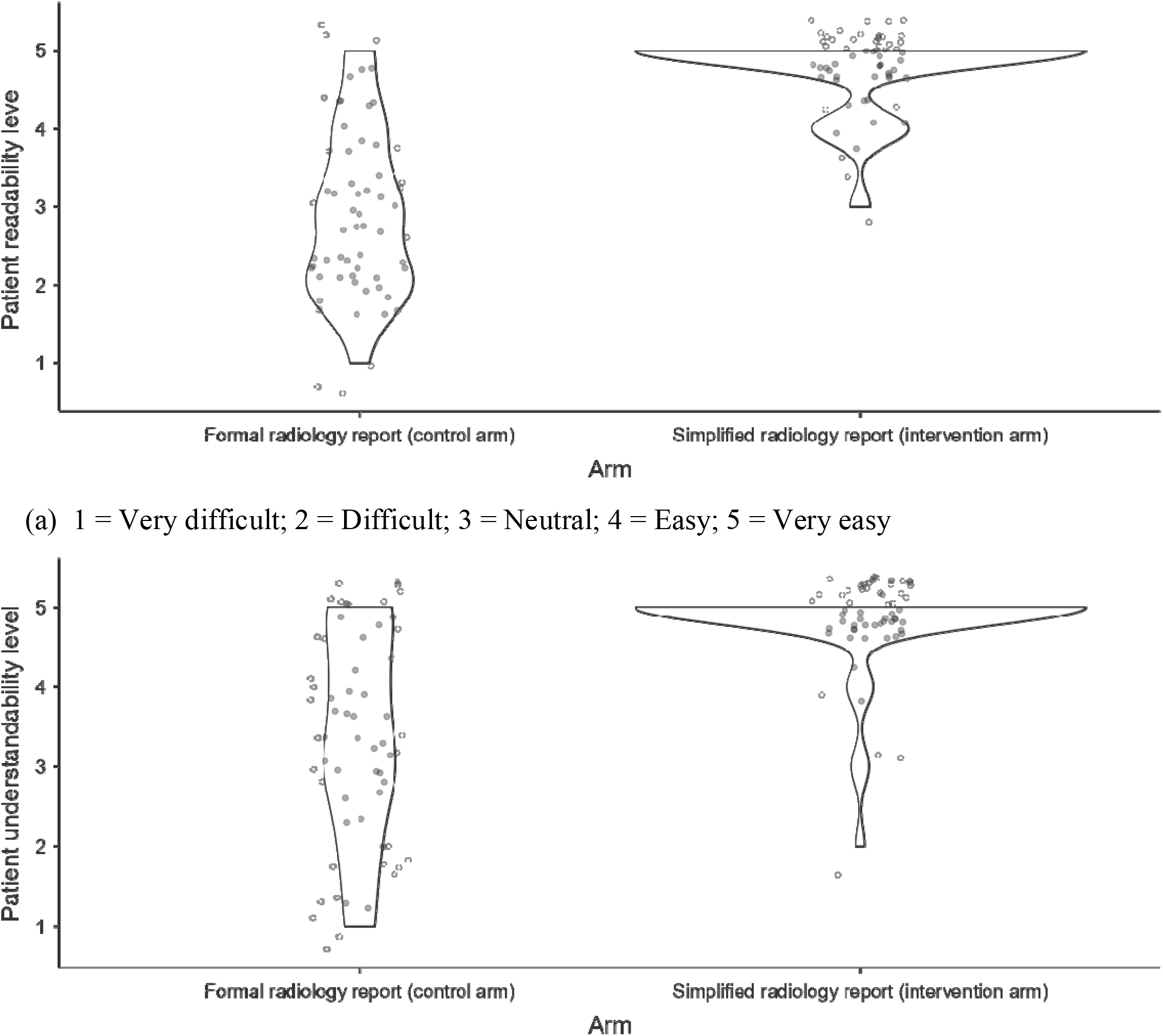
Subjective patient (a) readability and (b) understandability levels of both formal and GPT-4o simplified radiology reports within the intervention arm (Group B)

**Table 2:** Statistical comparison of the primary outcome.

|  | **Statistic\*** | **p-value** | **Effect size\*\*** |
| --- | --- | --- | --- |
| **Readability level** | 308 | <.001 | 0.83 |
| **Understandability level** | 613 | <.001 | 0.67 |

\*Mann-Whitney U test; \*\*Rank biserial correlation

The objective readability metric scores for both the formal and GPT-4o simplified reports are outlined in **Table 3**. The SMOG index, ARI and the Flesch-Kincaid Grade of the GPT-4o simplified reports received higher scores when compared with the formal reports within the intervention arm, whilst the Flesch Reading Ease score for the GPT-4o simplified reports had a lower score. These differences were statistically significant for all four indices. These score metrics are comparable to a 5th-9th grade reading level for the GPT-4o simplified reports, compared with the 11^th^-13^th^ grade reading level for the formal radiology reports.

**Table 3:** Statistical comparison of the objective readability scores.

|  | Formal report | Simplified report | Student's t | df* | p-value | Effect size |
| --- | --- | --- | --- | --- | --- | --- |
| Readability index | n = 59<sup>†</sup> | n = 59<sup>†</sup> |  |  |  |  |
| SMOG | 12.80 (2.15) [12.60] | 9.05 (1.16) [8.96] | 13.739 | 58 | <.001 | 1.79 |
| ARI | 10.33 (2.69) [10.34] | 7.12 (6.93) [6.35] | 3.43 | 58 | 0.001 | 0.447 |
| FRS | 26 (13) [27] | 81 (5) [81] | -31.39 | 58 | <.001 | -4.087 |
| FK | 12.33 (2.28) [12.60] | 5.79 (0.93) [5.80] | 22.89 | 58 | <.001 | 2.980 |

<sup>†</sup> Mean (SD) [Median]. \*Degrees of freedom. SMOG: Simple Measure of Gobbledygook Index, ARI: Automated Readability Index, FRS: Flesch Reading Ease score, FK: Flesch-Kincaid Grade

#### Understandability scores

Among participants who reviewed the GPT-4o simplified radiology reports, 95% reported understanding greater than half of the report compared with only 46% of patients who read the formal radiology report **(Table 1, Figure 2b)**. The GPT-4o simplified report scored a significantly higher rank sum than the formal report, with a strong positive rank biserial correlation **(Table 2)**.

### Secondary outcomes

A total of 97% and 100% of the simplified reports were evaluated as either ‘accurate’ or ‘very accurate’ by the first and second radiologists, respectively **(Table 4)**. The X^2^ test estimated a significant difference in the evaluation of factual accuracy between the two radiologists. Each radiologist identified hallucinations in only one of the simplified reports, with no statistically significant difference. None of the reported hallucinations was evaluated by either radiologist as being potentially harmful to the patient.

**Table 4:**
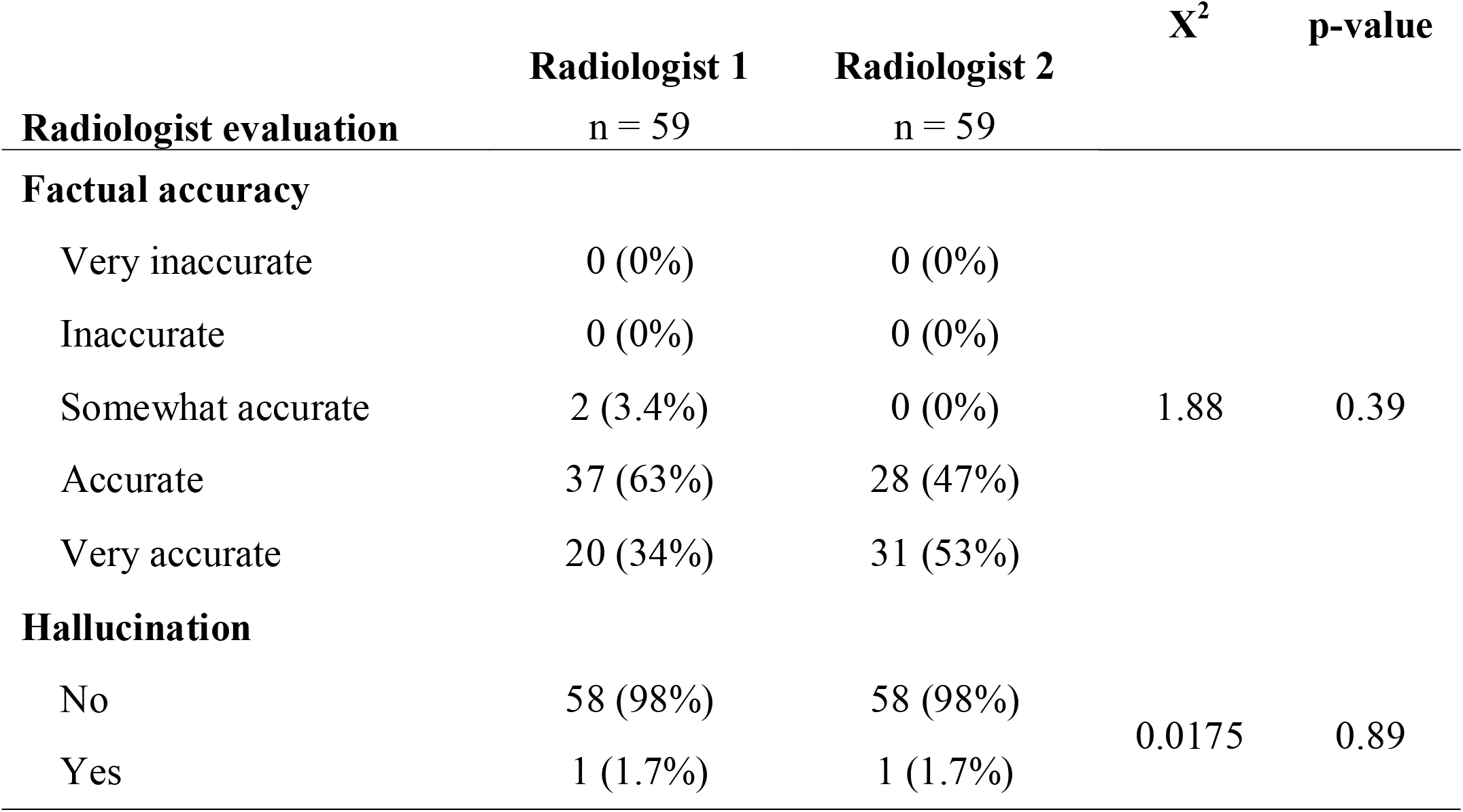
Statistical comparison of factual accuracy and hallucination.

Almost all participants (98.3%) indicated a preference to receive a simplified radiology report alongside the formal report in the future.

## Discussion

Our results indicate that LLM-simplified radiology reports improve patient readability and understandability whilst maintaining a high level of factual accuracy with minimal hallucination rates. This study is unique because each participant prospectively evaluated a simplified version of their own radiology report rather than a fictitious sample report, thereby ensuring that assessments were interpreted in a personalised, real-world clinical context. The strong participant (98.3%) demand for simplified reports to be provided alongside formal reports in the future demonstrates a clear appetite for patient-centric radiology reports. These findings are of utmost clinical significance in the current healthcare landscape, whereby widespread adoption of immediate patient access to electronic health records, including radiology reports, is increasingly common [1].

Despite improved patient access and transparency, radiology reports remain inherently challenging for patients to read and interpret, largely due to the use of medical jargon and a reporting style optimised for referring clinicians rather than a patient-centric focus. Objective readability metrics, including those utilised within this study, are derived from a combination of sentence length, word complexity and the number of syllables within each word. Although our study demonstrated that GPT-4o simplification successfully improved readability scores across all indices, the SMOG index and ARI did not meet the recommended equivalent to a sixth-grade reading level. The presence of lengthy, polysyllabic medical terminology in radiology reports likely contributed to the higher-than-expected SMOG index and ARI score, as these indices are particularly sensitive to syllabic complexity and word length, respectively. Therefore, the overall trend across multiple readability metrics rather than any single index provides a more reliable indicator of report readability. It is the authors’ view that oversimplification of radiology reports may lead to loss of essential clinical nuance and thus, efforts to improve readability must be balanced against the need to preserve diagnostic accuracy. Whilst technical jargon remains necessary to ensure accurate, standardised communication between radiologists and healthcare professionals to aid clinical decision-making, this often inadvertently harms patients by creating a complex imbalance between report accessibility and the health literacy required to read and comprehend radiology reports. This may lead to increased patient anxiety, stress or misinterpretation of the report findings [3, 4]. Bridging this health literacy gap is therefore critical not only to empower patients to assume a more active role in their own health care to make informed medical decisions, but also to foster a more collaborative relationship with their healthcare practitioner.

In Australia, recent legislative reform to MHR has either removed or reduced the seven-day delay for online patient access, with patients now being able to view X-ray reports immediately and other diagnostic imaging modalities following a five-day interval [2]. This legislative change further accentuates the risk of patient confusion and misunderstanding, increasing the likelihood that patients will access complex radiological findings before discussing them with their referring clinicians. An unintended consequence of this may be higher volumes of patient-initiated enquiries directed at clinicians in a healthcare system that is already operating at or near capacity. Notably, radiologists face significant workflow pressures, including an ever-increasing workload burden, disruptions, and burnout; therefore, it is imperative that any report simplification tool aimed at improving radiologist-patient communication should avoid exacerbating these demands [14]. This ultimately requires seamless integration of report simplification tools into existing radiological practice infrastructure. LLMs demonstrate promising potential to achieve this balance, with our study showing that LLMs can accurately simplify individualised radiology reports to a level that is both readable and comprehensible to patients within the relevant clinical context.

LLM performance is powerfully shaped by a plethora of factors, including prompt quality, contextual detail, task complexity and the volume of pre-trained data. In general, a more detailed prompt yields a more accurate and reliable text output. Prucker et al. utilised a single-sentence prompt, reporting a factual inaccuracy rate of 6% [24]. This contrasts with our study findings, which demonstrated that all GPT-4o simplified reports were rated as at least somewhat accurate by both radiologists and, significantly, did not raise concerns that would render its clinical use unsafe. This slight but notable discrepancy in factual accuracy across the studies may be partly attributable to differences in model architecture, radiologist subjectivity, and our use of a more specific and detailed zero-shot prompt. While increasingly sophisticated engineered prompts have been proposed to enhance LLM performance, our findings suggest that a detailed zero-shot prompt provides a practical and accessible approach for everyday patients without compromising report accuracy [25, 26].

A key barrier to integrating LLMs into clinical practice is the risk of hallucination, a phenomenon in which models generate plausible but factually incorrect or misleading information. This remains problematic within the radiology field and broader healthcare setting as this may again cause further misinterpretation of report findings, patient anxiety or false assurance in the event of a sinister pathology being portrayed as benign. Our results demonstrate minimal hallucination (1.7%) with each radiologist identifying only a single instance. Whilst hallucinations are likely an innate attribute of LLMs that cannot be eliminated, our results are concordant with the literature, which reports hallucination rates of up to 7%, with their existence highlighting the need for ongoing human oversight [16, 17, 19, 27]. Reassuringly, none of the hallucinations in our study resulted in patient harm, an observation that warrants vital consideration when assessing the viability of safe LLM integration into future radiological workflows. However, these findings are limited to our study cohort, and future research is required to validate these observations and establish guidelines defining acceptable thresholds for hallucination rates.

Patient data and privacy security remain at the forefront of any future efforts to integrate LLMs into clinical workflow. This study was approved by our institution’s Clinical Informatics Governance Committee, with all inputted de-identified radiology reports retained within the secure local IT network and in compliance with Australian privacy and security frameworks. This ensures that patient data is not available to train, retrain or improve the existing LLM. However, expanding the use of LLMs across national or global platforms, including MHR, would strain our local network infrastructure and require significantly more robust consumer data and privacy protections. Implementation at this scale must therefore comply with national privacy, cybersecurity and governance frameworks to ensure the secure handling and storage of sensitive patient data.

Several limitations were identified in our study. The main limitation is its single-centre study design, which limits the ability to extrapolate the findings to other patient cohorts. Similarly, our findings cannot be generalised to diagnostic imaging modalities not included in our research, such as magnetic resonance imaging or nuclear medicine scans due to their inherent complexity and more nuanced radiological interpretation. Participant responses may also be influenced by response bias as patients were able to distinguish the formal radiology report from the GPT-4o simplified version. This may result in the LLM-simplified reports having more favourable readability and comprehension scores.

## Conclusion

In summary, our study demonstrates that LLMs can accurately simplify radiology reports to a readability and comprehensibility level more appropriate for patients with minimal hallucination rates and without loss of clinical nuance. Whilst the presence of hallucinations reinforces the need for ongoing human oversight, the hallucinations encountered within our study did not critically result in patient harm. Therefore, LLMs show promising potential to enhance radiologist-patient communication without significantly impacting workflow, in an era of increasing patient access and demand for electronic health records. Future research on integrating LLMs into online patient portals, such as MHR, and on the potential data privacy pitfalls is required for successful implementation.

## Data Availability

All data produced in the present study are available upon reasonable request to the authors

## Acknowledgements

None.

